# Explainable advanced electrocardiography predicts coronary artery disease on coronary computed tomography angiography

**DOI:** 10.64898/2026.02.21.26346770

**Authors:** Manoj Rajamohan, Daniel E Loewenstein, Maren Maanja, Zaidon Al-Falahi, Abhinayeni Kuhasri, Kevin Yang, Chosita Cheepvasarach, Thomas Lindow, Todd T Schlegel, Yalu Wen, Patrick A Gladding, Martin Ugander, Rebecca Kozor

## Abstract

**BACKGROUND:** Conventional electrocardiography (ECG) has limited diagnostic accuracy for detecting coronary artery disease (CAD) in patients with stable chest pain. Advanced electrocardiography (A-ECG) may improve diagnostic performance. The study aimed to derive, externally validate, and prognostically validate an explainable A-ECG score for detecting CAD on coronary computed tomography angiography (CCTA).

**METHODS:** Participants attending an outpatient rapid access chest pain clinic (RACC) underwent a standard 12-lead ECG and CCTA. Any CAD was defined as any calcified or non-calcified plaque. Elastic net with nested resampling was used to derive an A-ECG score using measures from the conventional ECG, derived vectorcardiography, and measures of waveform complexity.

**RESULTS:** In the derivation cohort (n=171, age 59±13 years, 60% male), n=99 (58%) had any CAD on CCTA. A seven parameter A-ECG score to detect any CAD was derived. In an external validation cohort (n=773, age 57±12 years, 49% male, n=433 (56%) with any CAD), the score had an area under the receiver operating characteristic curve [95% confidence interval] of 0.66 [0.63–0.70] for detecting any CAD, and 0.72 [0.68–0.76] for detecting any coronary artery calcification. In the UK Biobank (n=27,239, 966 events, follow-up 1.9 [0.7–4.4] years, age 66±8 years, 50% female), higher A-ECG scores were associated with cardiovascular events even after adjusting for age, sex and cardiovascular risk factors (p<0.001).

**CONCLUSIONS:** An explainable A-ECG model, incorporating demographic and electrocardiographic features, demonstrated modest but externally reproducible discrimination for CCTA-defined coronary atherosclerosis and independent prognostic association in a large population cohort. This scalable, low-cost approach may aid triage and risk stratification in chest pain pathways.

## Introduction

Chest pain is one of the most common presentations to emergency departments with substantial cost associated with admission to hospital for further diagnostic assessment and treatment (1). By obviating the need for hospitalisation, the rapid access chest pain clinic (RACC) model has been shown to be a valuable outpatient clinical pathway for the safe and efficient assessment, diagnosis and treatment of patients presenting with symptoms suspicious of cardiac chest pain and at low to intermediate risk (2, 3). A RACC seeks to reduce the financial burden associated with unnecessary hospital admission for management of patients with low to intermediate risk chest pain, through the use of risk stratification algorithms such as the HEART score (4) and highly accurate diagnostic tests including coronary computed tomography angiography (CCTA)(5).

Standard 12-lead electrocardiography (ECG) is the initial diagnostic test used in the risk stratification of chest pain. However, conventional analysis of the ECG has a relatively low sensitivity and specificity for the diagnosis of clinically significant coronary artery disease (CAD) outside of the diagnosis of myocardial infarction (6, 7). The use and interpretation of the ECG have been largely unchanged in the past few decades. However, recent improvements in digital technology, specifically in signal processing and biomedical computing, have enabled newer innovative techniques of ECG analysis which have demonstrated promise, collectively termed advanced electrocardiography (A-ECG).

A-ECG refers to the integration of conventional ECG parameters with other ECG analysis techniques including, singular value decomposition measures of waveform complexity and derived vectorcardiographic (VCG) measures. All these measures are quantified from the standard resting 12-lead ECG. In clinical practice, A-ECG analysis can be performed on digitally acquired ECG files, both rapidly and inexpensively (8). No explainable ECG model has been validated against CCTA-defined plaque burden in contemporary chest pain pathways.

We hypothesised that A-ECG can improve the diagnostic assessment of intermediate risk chest pain in the RACC. The aim of our study was to derive and validate an A-ECG score for any CAD as detected by CCTA in a RACC patient population, and determine its association with prognosis.

## Methods

### Study Design

This was a prospective study to derive and validate an A-ECG score for the detection of any CAD, as defined by the presence of any calcified or non-calcified plaque or luminal stenosis in any epicardial coronary artery on CCTA. The derivation cohort consisted of patients who were prospectively referred for assessment at an academic hospital RACC between February 2017 and January 2022. External validation was subsequently performed in an independent external validation cohort drawn retrospectively from a separate healthcare institution, as well as an additionally separate prognostic cohort. The study was approved by local ethics committees and all participants either provided written informed consent or a retrospective waiver of consent was obtained.

### Derivation cohort

The derivation cohort inclusion criteria were age >18 years, referred to the outpatient RACC at Royal North Shore Hospital (Sydney, Australia) following presentation to the emergency department and assessed between February 2017 and January 2022, and underwent digitally recorded ECG and CCTA as part of the chest pain pathway work-up. Exclusion criteria were patients with known prior CAD, prior acute myocardial infarction, and previous revascularisation, including percutaneous coronary intervention and coronary artery bypass grafting.

### Validation cohort

The external validation cohort consisted of 773 patients (379 [49%] male, age 57±12 years) from North Shore and Waitakere Hospitals, Te Whatu Ora Waitematā Health, New Zealand, having undergone CCTA and ECG for assessment of CAD in setting of chest pain. For analysis of presence or absence of coronary calcification, the cohort consisted of 711 patients (356 [50%] male, age 57±12 years) due to the absence of non-contrast CT scan data for a minority (8%) of patients.

### Prognosis cohort

The prognostic performance of the A-ECG score for detecting any CAD was evaluated in a cohort from the United Kingdom (UK) Biobank – a large-scale database of volunteer participants from the UK(9). The prognostic endpoint was defined as a composite of the time to the first cardiovascular event or all-cause mortality following the index ECG. Baseline data collection for all subjects included a resting 12-lead ECG and an extensive questionnaire capturing demographic, clinical, family, and psychosocial parameters. For analysis, participants were divided into those without prior cardiovascular disease (CVD) and those with a documented or self-reported history of CVD, such as stroke, myocardial infarction, or angina. Subjects were excluded based on ECG criteria that may limit analysis, including atrial or ventricular arrhythmia or QRS duration >120 ms.

### ECG acquisition and analysis

Resting 12-lead ECG data for each subject were collected from the local ECG storage systems and exported into xml files. A-ECG semi-automatic software developed in-house was used to analyse the xml files. A-ECG analysis encompassed: conventional ECG measures derived automatically, including all major waveform intervals, axes and voltages; A-ECG measures, obtained after ECG signal averaging comprised of vectorcardiographic measures obtained from the Frank X, Y and Z leads derived from the 12-lead ECG using Kors’ regression transformation(10), and QRS and T-wave complexity measures quantified from eigenvector lead waveforms calculated using singular value decomposition of the 12-lead ECG(8, 11). Further exclusion criteria included non-sinus rhythm, bundle branch block (QRS>120ms), pre-excitation, heart rate > 100bpm, and corrupted or uninterpretable ECG files.

### CCTA analysis

Each clinically acquired CCTA was analysed by a study investigator (MR) blinded to the A-ECG results for the presence, distribution, and severity of CAD. Any coronary artery disease was defined as the presence of any coronary plaque (calcified or non-calcified) or luminal stenosis. Significant CAD was defined as luminal stenosis >50% in any coronary artery.

### Statistical Analysis

Statistical analysis was performed in R (R Foundation for Statistical Computing, version 4.1.2, Vienna, Austria). Continuous variables are reported as mean±SD and count data as number and frequency. Baseline characteristics were compared using the t test, or Mann-Whitney U-test, as appropriate depending on normal distribution. A multivariable A-ECG score for detecting any CAD was derived using elastic net logistic regression. Nested resampling was employed for model evaluation to determine cross-validated diagnostic accuracy and derive 95% confidence intervals (CIs). This approach was specifically selected to mitigate the risk of overfitting and minimise optimisation bias within the predictive model. The derived A-ECG score was then used to estimate the probability of a given subject in the validation cohort having any CAD. Prognosis was evaluated by Kaplan-Meier. Multivariable cubic spline analysis was generated to demonstrate changes in hazard ratio with increasing AECG score as a continuous variable, with adjustment of age, sex, hypertension, diabetes mellitus, dyslipidaemia, family history, smoking, and elevated body mass index.

## Results

### Study population

The final analysis of the derivation population included 171 patients (60% male, age 59±13 years). Table 1 summarises the baseline patient characteristics. Of these, 72 (42%) had normal coronary arteries and 99 (58%) had any CAD. The total cohort was deemed low-to-intermediate risk for major cardiac events as indicated by a HEART score of 3±1 (range 1-6). Any CAD group had a higher risk score compared to the normal coronaries group (HEART score, p<0.001). The any CAD group was older (p<0.001) and had a higher proportion of males (p=0.04). They were also more likely to be hypertensive (p=0.001), hyperlipidaemic (p=0.001) and smokers (p=0.02). Rates of diabetes did not differ between the two groups, and both groups were overweight as indicated by body mass index (BMI) (p=0.46). Table 2 summarises the patient characteristics of the validation cohort.

**Table 1:** Baseline patient characteristics of the derivation cohort.

| Parameters | All | Normal | Any coronary | p-value |
| --- | --- | --- | --- | --- |

|  |  | **coronaries** | **disease** |  |
| --- | --- | --- | --- | --- |
| Number of patients, n (%) | 171 (100) | 72 (42) | 99 (58) |  |
| Age, years | 59 ± 13 | 51 ± 12 | 64 ± 12 | <0.001 |
| Male sex, n (%) | 103 (60) | 37 (51) | 66 (67) | 0.04 |
| HEART score | 3 ± 1 | 3 ± 1 | 4 ± 1 | <0.001 |
| Body mass index, kg/m<sup>2</sup> | 27 ± 5 | 27 ± 5 | 28 ± 6 | 0.46 |
| Hypertension, n (%) | 62 (36) | 16 (22) | 46 (46) | 0.001 |
| Diabetes, n (%) | 18 (11) | 4 (6) | 14 (14) | 0.1 |
| Smoker, n (%) | 60 (35) | 18 (25) | 42 (42) | 0.02 |
| Hyperlipidaemia, n (%) | 77 (45) | 19 (26) | 58 (59) | <0.001 |

**Table 2:** Patient characteristics of the validation cohort.

| **Parameters** | **All** | **Normal coronaries** | **Any coronary disease** | **p-value** |
| --- | --- | --- | --- | --- |
| Number of patients, n (%) | 773 (100) | 340 (44) | 433 (56) |  |
| Age (mean (SD)) | 57 ± 12 | 53 ± 14 | 61 ± 10 | <0.001 |
| Male sex, n (%) | 379 (49) | 139 (41) | 240 (55) | <0.001 |
| Body mass index, kg/m<sup>2</sup> | 29 ± 6 | 29 ± 7 | 29 ± 6 | 0.5 |
| Hypertension, n (%) | 279 (36) | 81 (24) | 198 (46) | <0.001 |
| Diabetes, n (%) | 116 (15) | 42 (12) | 74 (17) | 0.06 |
| Hyperlipidaemia, n (%) | 108 (14) | 37 (11) | 71 (17) | 0.05 |

### Advanced ECG analysis

In the derivation analysis, A-ECG analysis of the derivation cohort identified seven measures that had the strongest association with any CAD and included: (1) Increasing age, (2) the percent of the total QRS loop area in the frontal plane that subtends the left lower quadrant of that frontal plane, (3) the elevation angle (degrees) of the 3-dimensional QRS loop when three eighths of the way into the loop, (4) the elevation angle (degrees) of the 3-dimensional QRS loop when six eighths of the way into the loop, as the ST segment loop extends from the end of the QRS wave to the end of the T wave (5) T-wave complexity, defined as the sum of T-wave eigenvectors 3 to 8 divided by (eigenvector 1 minus eigenvector 2), (6) voltage sum of T-wave eigenvectors 4 to 8, and (7) sex.

These features were combined to derive a seven parameter A-ECG score to detect any CAD with an area under the receiver operating characteristic (ROC) curve of [95% confidence interval] of 0.78 [0.77–0.78], sensitivity 82 [81–83]%, specificity 54 [53-55]%, positive predictive value 71 [71–72]%, negative predictive value 70 [68–71]%, positive likelihood ratio 1.8 and inverse negative likelihood ratio 3.0. The measures included in the final seven-parameter A-ECG score, and the intercept and coefficients for the regression equations are presented in Tables 3 and 4. Diagnostic performance measures are shown in Table 5.

**Table 3:** A-ECG & clinical measures in the score.

|  | **Total (n=171)** | **Normal coronaries (n=72)** | **Any stenosis (n=99)** | **p-value** |
| --- | --- | --- | --- | --- |
| Age | 62.1 (52.8-73.1) | 54.7 (44.3-62.5) | 69.2 (60.3-76.3) | <0.001 |
| Total QRS loop area in the frontal plane subtending the left lower quadrant, (%) | 0.90 (0.05-5.4) | 0.94 (0.05-7.1) | 0.89 (0.06-4.7) | 0.01 |
| Elevation angle of the 3D QRS loop when 3/8 of the way into the loop, degrees | 28.8 (15.4-38.9) | 33.1 (20.3-41.4) | 26.2 (13.4-35.5) | <0.001 |
| Elevation angle of the 3D QRS loop when 6/8 of the way into the loop, as the ST segment loop extends from the end of the QRS to end of the T-wave, degrees | 31.9 (25.1-41.2) | 35.8 (28.8-44.6) | 29.8 (19.7-38.9) | 0.004 |
| T-wave complexity, defined as the sum of T-wave eigenvectors 3 to 8 divided by (eigenvector 1 minus eigenvector 2), unitless ratio | 0.09 (0.06-0.16) | 0.09 (0.06-0.14) | 0.10 (0.07-0.21) | 0.02 |
| voltage sum of T-wave eigenvectors 4 to 8, mV | 17.8 (15.1-20.9) | 16.8 (14.3-19.7) | 18.5 (15.6-21.1) | 0.01 |
| Male sex, n (%) | 103 (60) | 37 (51) | 66 (67) | 0.04 |

**Table 4:** A-ECG score component estimates.

| **Term** | **Estimate** | **Standardized coefficient** |
| --- | --- | --- |
| Age | -0.05681443 | -0.754 |
| Male sex | -0.1892255 | -0.093 |
| Total QRS loop area in the frontal plane subtending the left lower quadrant | 0.004451245 | 0.033 |
| Elevation angle of the 3D QRS loop when 6/8 of the way into the loop, as the ST segment loop extends from the end of the QRS to end of the T-wave | 0.001766589 | 0.030 |
| T-wave complexity, defined as the sum of T-wave eigenvectors 3 to 8 divided by (eigenvector 1 minus eigenvector 2) | -0.1309184 | -0.029 |
| Elevation angle of the 3D QRS loop when 3/8 of the way into the loop | 0.000747439 | 0.015 |
| Voltage sum of T-wave eigenvectors 4 to 8 | -0.00208173 | -0.011 |
| (Intercept) | 3.39465903 | 0 |

**Table 5:** Performance markers in the A-ECG score to detect any CAD or any CAC.

| **Performance Marker** | **Derivation cohort** | **Validation cohort**<br>**Any CAD** | **Validation cohort**<br>**Any CAC** |
| --- | --- | --- | --- |
| AUC (95% CI) | 0.78 [0.77–0.78] | 0.66 [0.62–0.70] | 0.72 [0.68–0.76] |
| Sensitivity, % | 82 [81–83]% | 90 [88–92]% | 91 [90–93]% |
| Specificity, % | 54 [53–55]% | 31 [29–33]% | 35 [33–37]% |
| Positive predictive value, % | 71 [71–72]% | 63 [61–64]% | 68 [66–69]% |
| Negative predictive value, % | 70 [68–71]%, | 70 [68–71]% | 73 [71–75]% |
| Positive likelihood ratio | 1.8 | 1.3 | 1.4 |
| Inverse Negative likelihood ratio | 3.0 | 3.1 | 4.0 |

Abbreviations: A-ECG, advanced electrocardiography; CAD, coronary artery disease; CAC, coronary artery calcification; AUC, area under the curve; CI, confidence interval

In the validation analysis, sensitivity analyses were undertaken in the validation cohort. Diagnostic performance measures for the derivation and validation cohorts are shown in Table 4. In the validation cohort, the A-ECG score’s diagnostic utility was lower for the detection of any CAD. The sensitivity showed a substantial numerical increase, whilst specificity, positive predictive value and positive likelihood ratio were reduced. Negative predictive value and inverse negative likelihood ratio remained similar. Interestingly, the same score had a less attenuated diagnostic performance for detection of any coronary calcification. For this endpoint, reduction in specificity was offset by a substantial increase in sensitivity and inverse negative likelihood ratio – thereby retaining its ability to exclude calcified plaque.

In the prognosis analysis, the UK Biobank population for prognostic validation consisted of 27,239 subjects, mean age 66±8 years, 50% male. The mean follow-up duration was 1.9 [0.7-4.4] years with 966 cardiovascular events occurring over this period. Of these subjects, 23,051 (85%) had no previous cardiac history at the time of their inclusion. The baseline characteristics of the UK Biobank cohort are presented in Table 6. The A-ECG score is associated with prognosis in the form of cumulative rate of cardiovascular hospitalisation or all-cause death (figure 1) and hazard ratio for cardiovascular events adjusted for age, sex and cardiovascular risk factors (Figure 2).

**Figure 1:**
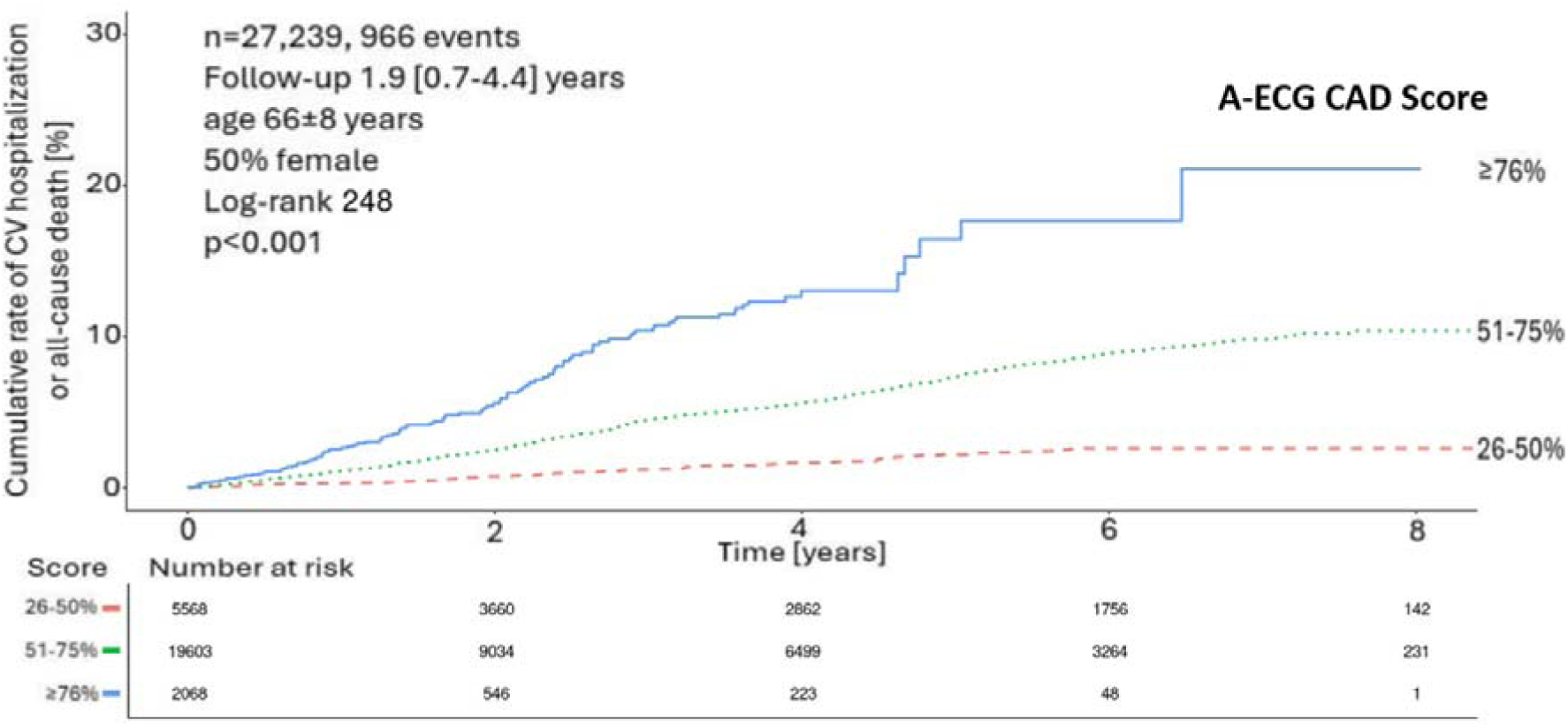
Cumulative rate of cardiovascular hospitalisation or all-cause mortality per quartile of A-ECG score. There were no subjects with a score ≤25%.

**Figure 2:**
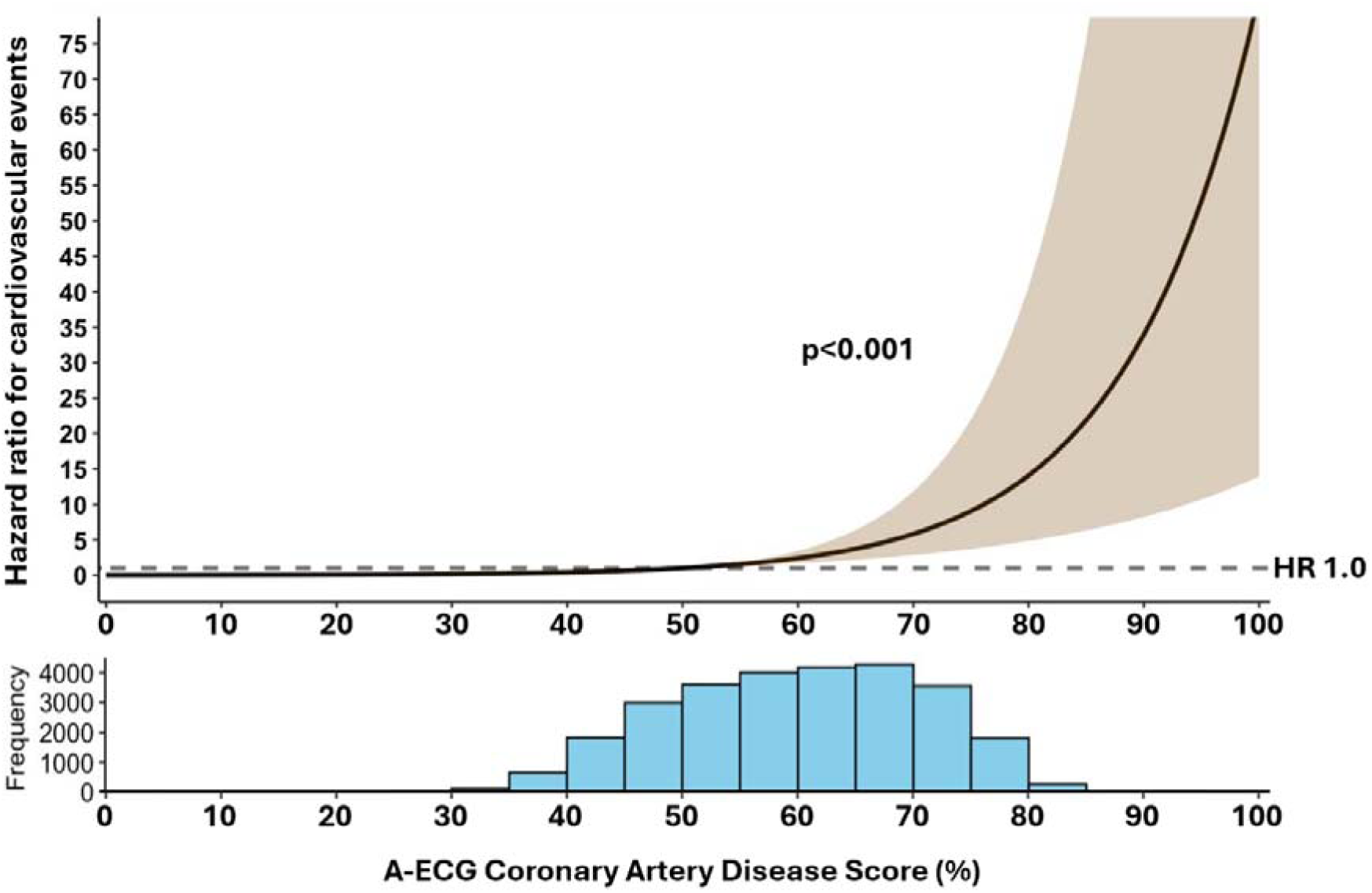
Restricted cubic spline of adjusted hazard ratio for cardiovascular events and all-cause mortality after adjustment for age, sex and cardiovascular risk factors.

**Table 6:**
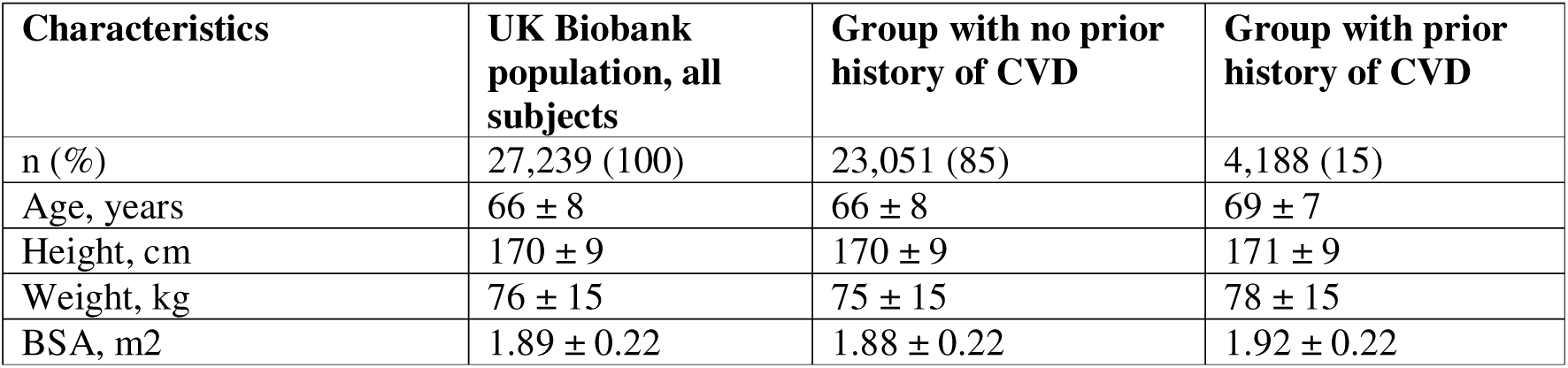
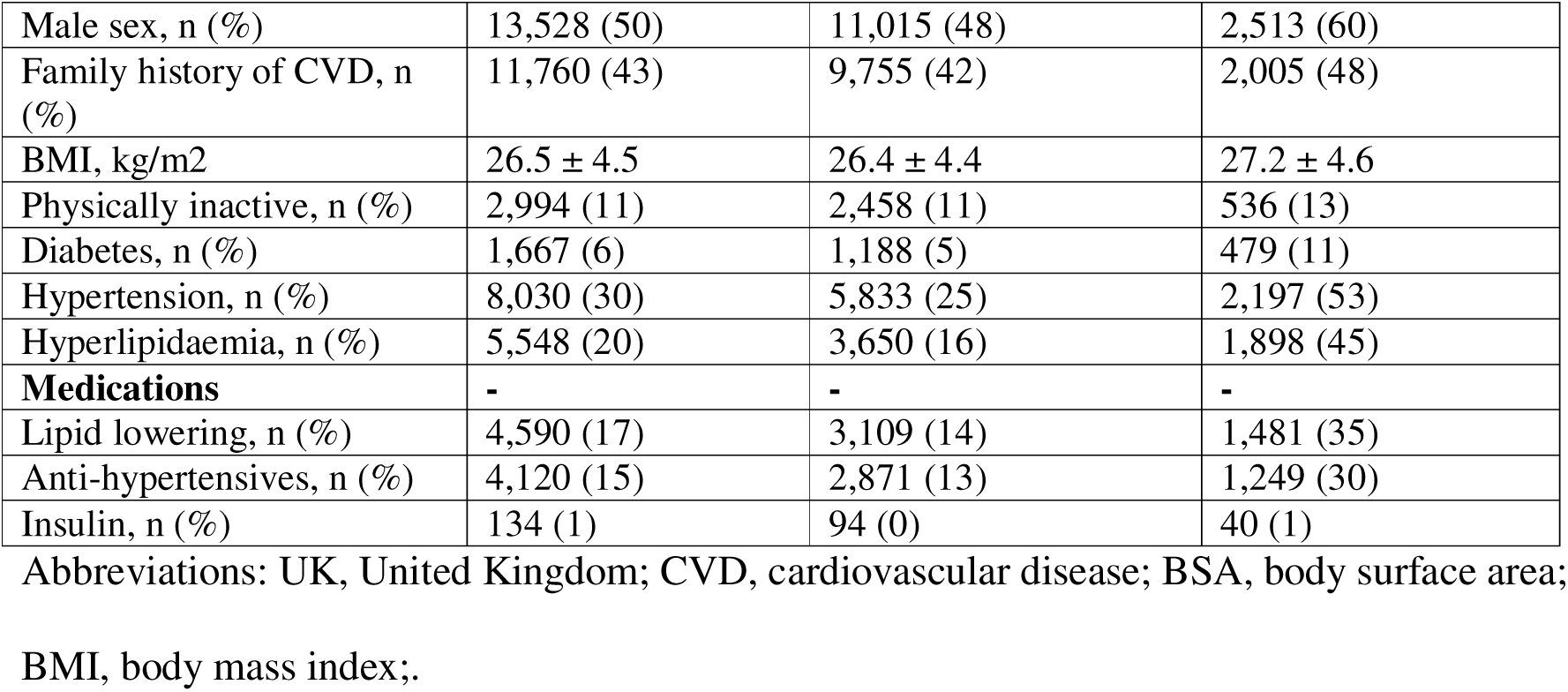
UK Biobank cohort patient characteristics.

## Discussion

The main finding of the study is that explainable A-ECG analysis of the resting standard 12-lead ECG can identify the presence of CAD on CCTA with good diagnostic accuracy and an independent prognostic association. A seven-parameter model incorporating age, sex and five A-ECG measures achieved an AUC of 0.78 for detecting any CAD in a low-to-intermediate risk chest pain derivation cohort and an AUC 0.66 in an external validation cohort, while also achieving an AUC of 0.72 for detecting any coronary calcification in the validation cohort.

Importantly, the score also stratified long-term cardiovascular prognosis adjusted for age, sex and cardiovascular risk factor in a large external population cohort, supporting its clinical utility.

This is the first study to derive an explainable A-ECG score using CCTA-defined CAD, and it was done using a contemporary low-to-intermediate risk chest pain population. While previous investigations have shown that A-ECG improves the detection of catheterization-confirmed CAD and structural heart disease compared with conventional ECG interpretation, most have focused on invasive angiographic endpoints or functional ischemia. For example, prior studies have shown the ability of A-ECG to detect the presence of catheterization-proven or stress perfusion imaging-proven CAD with higher sensitivity and specificity than conventional ECG interpretation (8, 12). Additionally, other studies have demonstrated the utility of A-ECG in identifying left ventricular systolic dysfunction, left ventricular hypertrophy and other cardiac conditions (13–15). In contrast, the present study leverages CCTA as the reference standard, reflecting the current guideline-endorsed first-line imaging modality for stable chest pain evaluation (16–18). This is clinically important, as CCTA identifies both obstructive and non-obstructive plaque, which are increasingly recognised as prognostically meaningful.

### Clinical implications in the RACC setting

Patients presenting with low-to-intermediate risk chest pain represent a major diagnostic and resource burden. Although RACC pathways incorporating CCTA are cost-effective and safe, a substantial proportion of patients ultimately demonstrate no CAD, raising the need for upstream screening strategies to refine further testing referral. The present A-ECG score offers a rapid, low-cost, and scalable approach that could be applied at the point of first assessment.

Within the external validation cohort, an A-ECG score threshold of 50% yielded an inverse negative likelihood ratio of 3.1 for any CAD and 4.0 for any CAC, offering modest utility to justify no further testing or guide preventative pharmacotherapy respectively. Importantly, since the score is continuous, diagnostic certainty increases significantly as values deviate from 50%. Lowering the threshold to 40% increases the inverse negative likelihood ratio to 8.0, exponentially improving its discriminative power for CAD. This continuous feature allows clinicians to apply tailored thresholds and supports its potential role as an adjunct to clinical risk scores and screening, enabling more targeted use of CCTA or other imaging.

### Mechanistic insights from A-ECG features

The discriminative capacity and explainability of the A-ECG model is underpinned by physiologically meaningful features. Two of the seven measures reflect T-wave waveform complexity, a marker of ventricular repolarization heterogeneity. Increased T-wave complexity has consistently been associated with adverse cardiovascular outcomes and long-term mortality(19, 20), arrhythmic risk (increased risk for sudden cardiac death, appropriate therapies in patients with new ICD implants and identification of long QT syndrome compared to healthy controls(21, 22)), and has been shown to detect subclinical ischemia in asymptomatic populations undergoing exercise stress testing(23). Therefore, the inclusion of such T-wave measures support the concept that early ischemic or structural myocardial alterations manifest as subtle repolarization abnormalities, even in the absence of overt ECG changes.

Three further ECG measures relate to QRS loop orientation and spatial geometry, reflecting ventricular depolarization patterns. Alterations in QRS vector direction and loop morphology have long been associated with regional myocardial injury, ischemia, and adverse prognosis(24–31). Our findings align with prior work demonstrating that vectorcardiographic metrics provide incremental diagnostic information beyond conventional axis and voltage measurements, particularly in patients with functionally significant CAD or regional wall motion abnormalities(32, 33).

Finally, the inclusion of age and male sex reflects these well-established demographic risks for CAD(34, 35), and indeed they have the numerically strongest standardized coefficients in the score, indicating the relatively strongest contributions to the score. Their integration with electrocardiographic biomarkers enables a hybrid physiological-clinical risk model, which may enhance generalisability and interpretability. Importantly, the score retains a prognostic association despite adjusting for age and sex, confirming the incremental prognostic contribute of the A-ECG measures beyond age and sex alone.

### External validation and prognostic relevance

While specificity, positive predictive value and positive likelihood ratio declined in the external validation cohort, sensitivity improved. Importantly, AUC, negative predictive value and inverse negative likelihood ratio remained similar. This suggests that the model may preferentially exclude subclinical atherosclerotic disease in heterogeneous populations.

Notably, diagnostic performance was superior for detecting CAC. As calcification occurs later in the progression of CAD, it likely causes more extensive structural remodelling, resulting in more severe and easily detectable A-ECG abnormalities. Importantly, in the UK Biobank cohort, the A-ECG score was associated with cardiovascular events and mortality even after adjustment for age, sex and cardiovascular risk factors, indicating that the electrocardiographic signatures captured by the model reflect biologically meaningful myocardial substrate rather than purely anatomical plaque burden.

### Limitations

Several limitations merit consideration. The derivation cohort was modest in size and consisted mostly of non-obstructive CAD, reflecting real-world RACC populations, but limiting precision for detecting high-grade stenoses. However, nested resampling was employed to mitigate overfitting, and the performance was evaluated in a robustly sized validation cohort (n=773). CCTA was used as the reference standard, and therefore microvascular dysfunction or vasospastic disease may have been misclassified as “normal.” Finally, the study was observational and clinical utility will require larger-scale prospective studies.

## Conclusions

An explainable A-ECG model, incorporating demographic and electrocardiographic features, demonstrated modest but externally reproducible discrimination for CCTA-defined coronary atherosclerosis and independent prognostic association in a large population cohort. This approach provides an explainable and mechanistically interpretable, scalable, and low-cost screening tool that could complement existing clinical pathways by refining downstream testing selection. Prospective trials are warranted to evaluate its impact on diagnostic efficiency, cost-effectiveness, and patient outcomes.

## Data Availability

All data produced in the present study are available upon reasonable request to the authors.

